# Spatial assessment of stromal B cell aggregates predicts response to checkpoint inhibitors in unresectable melanoma

**DOI:** 10.1101/2024.08.09.24311758

**Authors:** James W. Smithy, Xiyu Peng, Fiona D. Ehrich, Andrea P. Moy, Mohammad Yosofvand, Colleen Maher, Nathaniel Aleynick, Rami Vanguri, Mingqiang Zhuang, Jasme Lee, MaryLena Bleile, Yanyun Li, Michael A. Postow, Katherine S. Panageas, Travis J. Hollmann, Margaret K. Callahan, Ronglai Shen

## Abstract

Quantitative assessment of multiplex immunofluorescence (mIF) data represents a powerful tool for immunotherapy biomarker discovery in melanoma and other solid tumors. In addition to providing detailed phenotypic information of immune cells of the tumor microenvironment, these datasets contain spatial information that can reveal biologically relevant interactions among cell types.

To assess quantitative mIF analysis as a platform for biomarker discovery, we used a 12-plex mIF panel to characterize tumor samples collected from 50 patients with melanoma prior to treatment with immune checkpoint inhibitors (ICI). Consistent with prior studies, we identified a strong association between stromal B cell percentage and response to ICI therapy. We then compared pathologist assessment of lymphoid aggregates with a density based clustering algorithm, DBSCAN, to both automatically detect B cell aggregates and quantify their size, morphology, and distance to tumor. Spatial neighborhood analysis identified TCF1+ and LAG3-T cell subpopulations enriched near stromal B cells. These analyses provide a roadmap for the further development and validation of spatial immunotherapy biomarkers in melanoma and other diseases.

## INTRODUCTION

As immune checkpoint inhibitors (ICIs) exert their anti-tumor efficacy by modulating complex interactions among multiple cell types in the tumor microenvironment (TME), the identification of predictive biomarkers for these agents in melanoma has been challenging. While useful in other tumor types, tumor-intrinsic features such as tumor mutational burden (TMB) and programmed death ligand 1 (PD-L1) have had limited clinical utility in melanoma in distinguishing patients most likely to respond to ICI.^1–3^ Initial translational studies characterizing the TME have suggested that more highly immune-infiltrated melanomas are more likely to benefit from ICI therapy--these include higher numbers of infiltrating CD8^+^ T cells,^4^ the presence of an interferon gamma transcriptional signature,^5^ and tumor expression of interferon gamma signaling components.^6^ Importantly, none of these reported biomarkers take spatial information into consideration, thus missing a critical feature of the TME.

Recent advances in multiplex spatial imaging technologies offer powerful tools for biomarker discovery by enabling the simultaneous detection of multiple markers of interest on a single tissue slide to delineate spatially oriented features of the TME.^7,8,9,10^ Among spatial technologies, multiplex immunohistochemistry/multiplex immunofluorescence (mIHC/mIF) assays are perhaps the most readily translatable into clinical practice, as robust IHC signals can be adapted to highly reproducible single-plex assays that can be run in clinical labs. The potential for these technologies across solid tumor oncology is broad— a recent meta-analysis involving over 10 different solid tumor types in 8135 patients demonstrated that mIHC/mIF was associated with improved performance in predicting response to PD-L1/ PD-1 treatment when compared to PD-L1 IHC, TMB, or gene expression profiling alone. ^11^

Integration of multiple cellular features with mIF/mIHC enables deeper investigation of the TME compared to single-plex assays,^9^ and the spatial relationships among cell types can reveal intercellular interactions with immunologic and clinical relevance.^12^ For instance, the spatial organization of PD1^+^ CD4^+^ T cells, tumor cells, and other immune cells was used to effectively predict response to pembrolizumab in cutaneous T-cell lymphoma, though there were no differences in pre-treatment cellular compositions by count alone.^13^ In melanoma, the spatial relationship between cytotoxic T cells and PD-L1 expressing macrophages was predictive of ICI response in a cohort of 14 melanoma patients.^14^ Among the most promising spatial biomarkers of ICI response are tertiary lymphoid structures (TLS), aggregates of B cells and other immune cell types that have been associated with ICI response in melanoma^15^ and other cancers.^16–18^ While the associations between TLS and ICI response are strong in these datasets, there are significant barriers to their widespread clinical application. Specifically, there is no consensus definition of TLS across studies, and their identification currently requires manual assessment of both phenotypic and spatial features by a trained pathologist. As such, automated methods to identify and quantify cellular features of these structures may provide a more standardized and efficient way to incorporate TLS into clinical practice.

To examine both the relative abundance and spatial organization of immune cell populations associated with response to ICI therapy in a cohort of patients with unresectable melanoma, we applied a 12-plex mIF panel to pre-treatment whole-tissue sections to identify features associated with response to immune checkpoint blockade We then leveraged a novel automated method for identification of stromal B cell aggregates and compared its effectiveness with pathologist assessments of lymphoid aggregates in predicting ICI response. Together, these data may inform the development of future tissue-based immunotherapy biomarkers in melanoma and other tumor types.

## RESULTS

### Patient & biospecimen characteristics

Fifty patients with unresectable melanoma and available archival pre-treatment tumor specimens were included in the primary analysis cohort. Patient demographics are summarized in **Supplementary Table 1.** Most patients were treated with anti-PD(L)1 monotherapy (58%), and the remainder were treated with combined PD-1 and CTLA-4 targeted therapy (36%) or CTLA-4 monotherapy (6%). Patients were treated with a median of 1 (interquartile range [IQR]: 0, 1) prior systemic therapy prior to study therapy; 40% of patients received a prior PD-(L)1 or CLTA-4 targeted ICI before study tissue was acquired.

Tissue specimens included 10 primary tumors (20%), 25 soft tissue and lymph node metastases (50%) and 15 visceral metastases (30%). The median time between biospecimen collection and start of study therapy was four months (IQR: 1, 8 months), and 40% of patients had non-PD-1-targeted intervening systemic therapy between tissue collection and the start of study therapy. All patients had progressed on prior therapies prior to staring study therapy.

### Immune populations associated with response to ICI therapy

Whole-tissue sections in the primary analysis cohort were stained with a 12-plex mIF panel, and over 20 million derivative cells were assigned to one of seven primary phenotypic cell types based on the combinatorial expression of multiple markers^19^ (**Figure 1A-E; STAR Methods**). These cell types were then projected onto a spatial pseudoplot for visualization (**Figure 1F**). The median number of total cells per case was 154,075 (IQR: 28,528, 566,764), with a median of 8,555 (IQR: 1,627, 56,205) immune cells per case. We analyzed cell populations within both the tumor and stromal compartments, comparing patients whose melanoma did and did not radiographically respond to study therapy.

**FIGURE 1.**
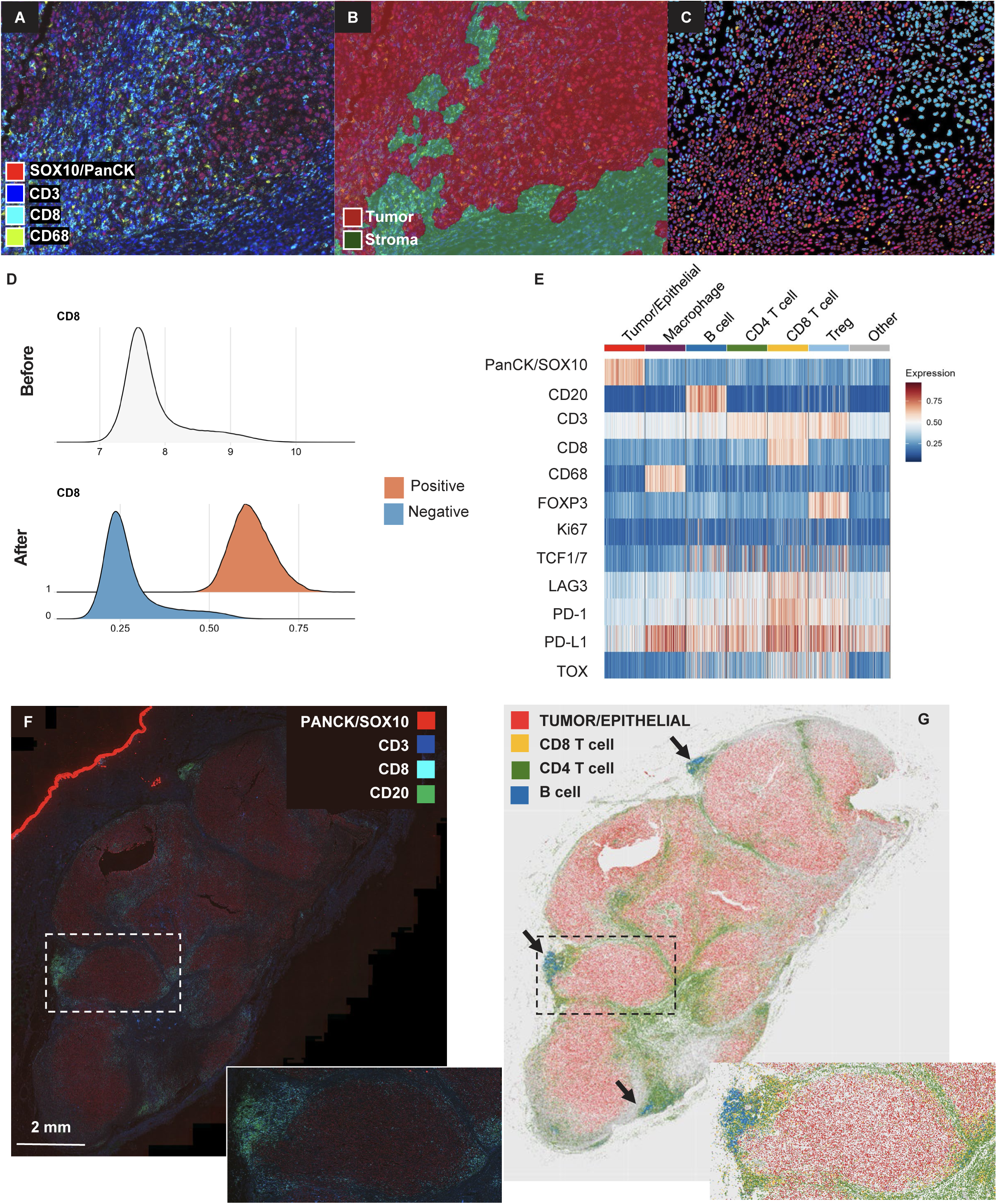
Multiplex immunofluorescence cell phenotyping pipeline. A. **Representative stacked TIFF images** demonstrating SOX10^+^ melanoma cells (red), CD68^+^ macrophages (yellow) and CD3^+^ T cells (dark blue), and CD8^+^ T cells (teal). B. **Representative tumor and stromal compartments**on melanoma whole-tissue sections, where identified using a self-supervised random forest classifier C. Representative nuclear and cytoplasmic segmentation identifying individual cells in the cell-mask digital output format. D. **Density plot of CD8**^+^ **signal intensity** for representative melanoma case, before (upper) and after (lower) binarization into positive and negative populations integrating both the visually identified threshold and average signal intensity on a cell-level basis. E. **Heatmap** demonstrating expression of 12 mIF markers across seven major cell types. F. **Merged mIF image** of a representative melanoma case with four channels: PANCK/SOX10 (red), CD3 (royal blue) CD8 (teal) CD20 (green). G. **Pseudoplot** of representative melanoma case demonstrating tumor/epithelial cells (red), CD8^+^ T cells (yellow), CD4^+^ T cells (green) and CD20^+^ B cells (blue). Arrows (black) indicate B cell aggregates.

While there were no significant differences in the absolute density of any cell population within the tumor compartment between responders and non-responders (**Supplementary Figure 1**), multiple CD20^+^ B cell populations in the stromal compartment were enriched in responders both as a fraction of all nucleated cells and as a fraction of immune cells (**Figure 2A**) compared to non-responders. A heatmap showing stromal percentages of cell populations in responders and non-responders is shown **Figure 2B** with denoted significance. Multiple immune cell types, including CD4^+^ and CD8^+^ T cell populations, are enriched among responding patients. The association of stromal CD20^+^ B cell percentage with radiographic response to ICI persisted even when the region of interest was confined to the stroma within 100 μm of the tumor boundary (**Supplementary Figure 2**). The percentage of stromal CD20 B cells among all nucleated cells effectively discriminated responders and non-responders with an area under the receiver operating characteristic curve (AUC) of 0.825 (**Figure 3A-C**). An optimal cutpoint of 0.644% for stromal B cell percentage in the total stroma was associated with improved progression-free survival (PFS) (p < 0.001; **Figure 3D**) and overall survival (OS) (p<0.001; **Figure 3E**). LDH levels and ICI treatment type did not significantly differ between patients with high and low stromal B cell percentages. M stage was associated with B cell percentage classification (p = 0.023), with the largest proportion of B cell high patients having M1b disease (40%) and the largest proportion of B cell low patients having M1c disease (57%). To avoid potential confounding by normal lymph node architecture, we repeated the analysis excluding 10 lymph node metastases. In this smaller cohort, the fraction of stromal CD20^+^ B cells remained an effective discriminator of response with an AUC = 0.772 (**Supplementary Figure S3**). The optimal cutpoint of 0.644% (determined in the full cohort) remained associated with improved PFS (p = 0.002) and OS (p = 0.009).

**FIGURE 2.**
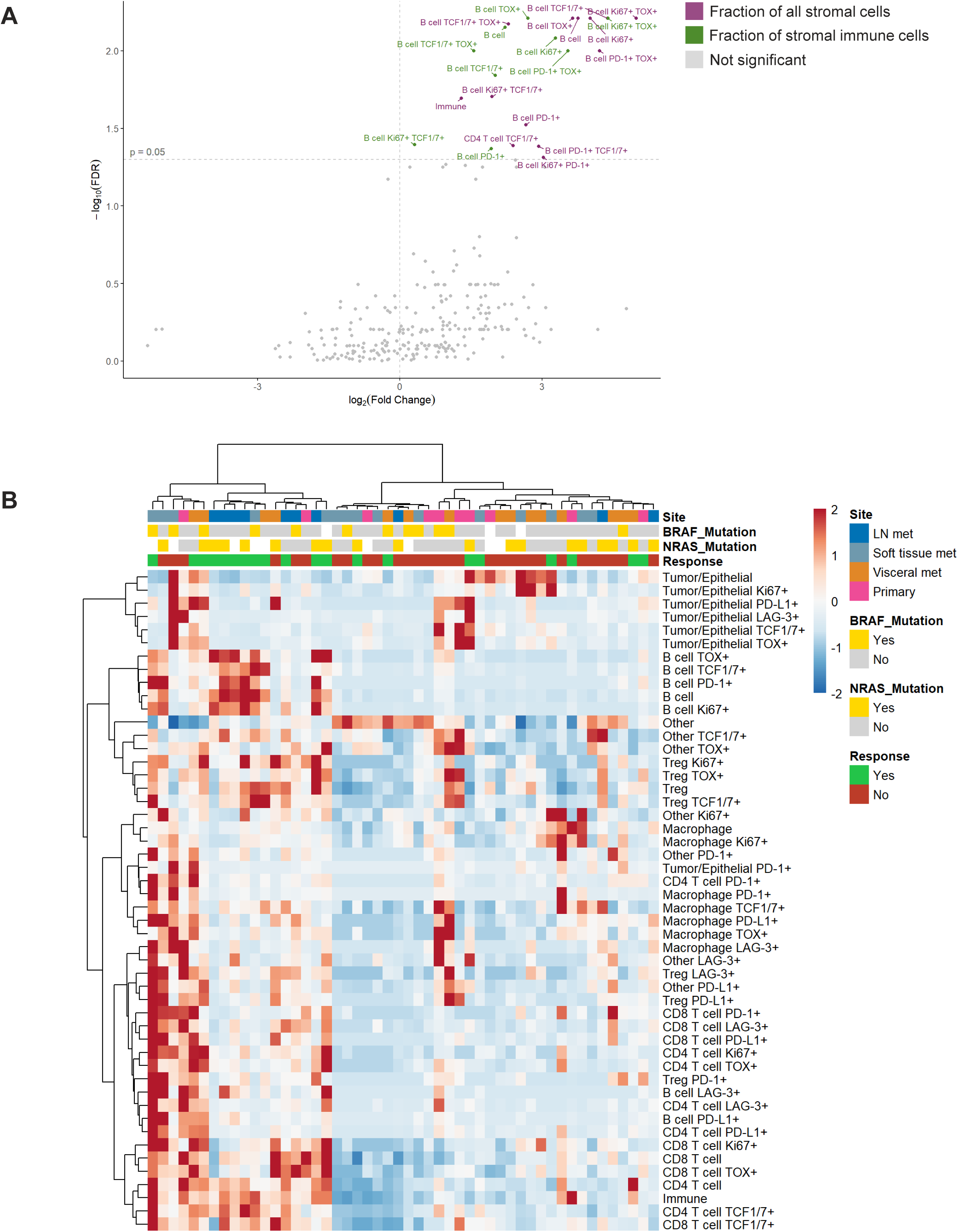
Stromal immune cells enriched in ICI-responsive melanoma samples. **A. Volcano plot of stromal immune cell populations** enriched in patients with complete or partial response to ICI therapy. Immune cell populations were calculated as a percentage of all stromal cells (purple) and as a fraction of stromal immune cells (green). **B. Heatmap of stromal percentages of cell populations and clinical characteristics.**A subset of the 265 cell populations is presented for visibility: major cell types, each major cell type expressing each functional marker, and total immune cells.

**FIGURE 3.**
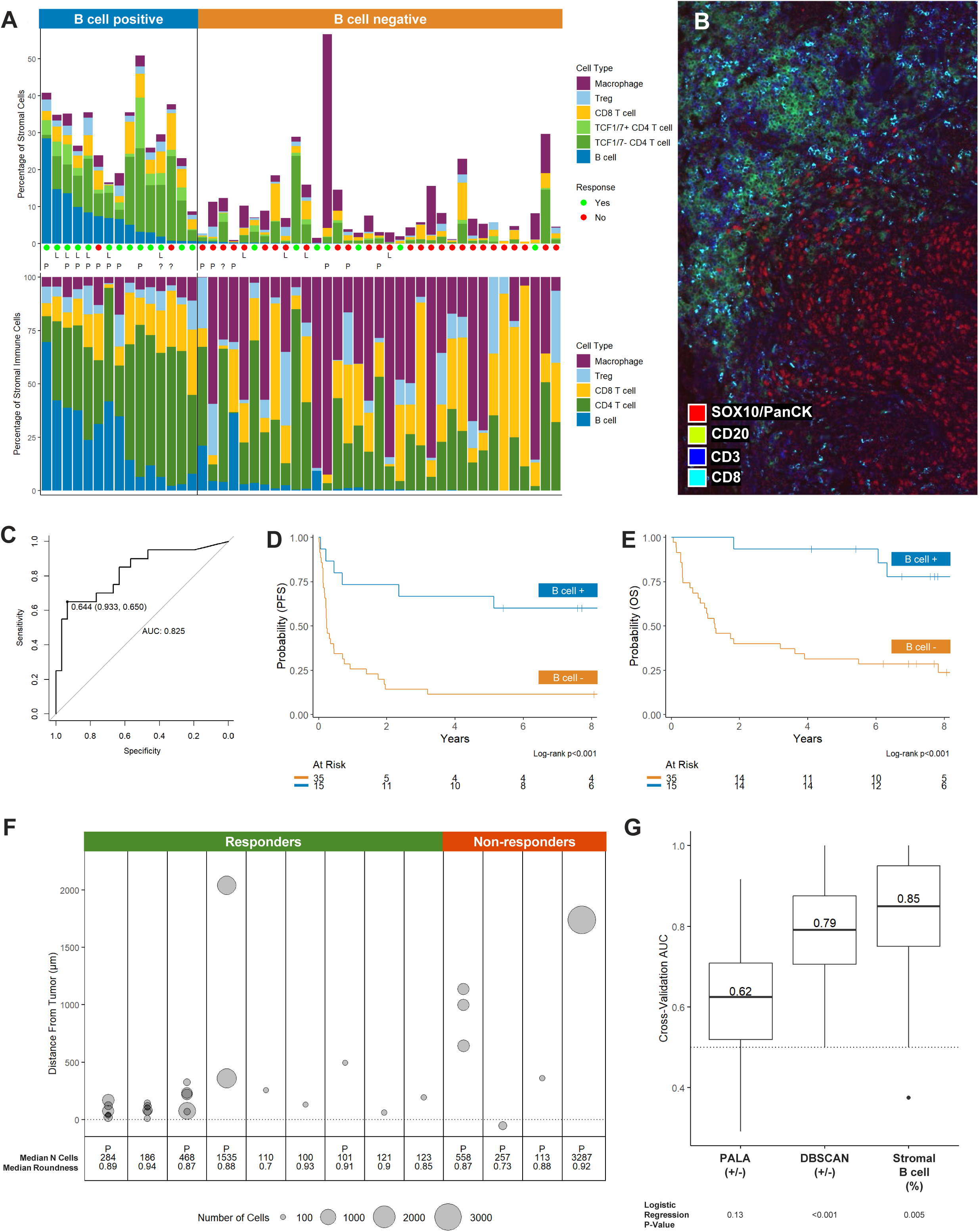
Quantitative assessment of B cells and association with clinical outcomes. **A. Bar plot of six key immune cell types, sorted by percentage of stromal B cells**. The top panel displays immune cells as a percentage of all stromal cells, and the bottom panel demonstrates relative populations as a fraction of all immune cells. Green circles indicate patients with radiographic PR or CR as their best response to ICI therapy. Red circles indicate patients with SD or PD to ICI therapy. “**L**” denotes lymph node specimen and “**P**” denotes presence of pathologist-assessed lymphoid aggregate (PALA). The vertical line indicates a stromal B cell percentage of 0.644% and separates B cell “positive” from B cell “negative” cases based on an optimal cutpoint. **B.** Representative image of a soft tissue a with high fraction of stromal B cells with B cell aggregate. **C. Receiver operating characteristic curve of** stromal B cell percentage in discriminating radiographic response to ICI therapy among all cases (n = 50). Area under the curve = 0.832; optimal cutpoint = 0.644%; **D. Progression-free survival** stratified by stromal B cell percentage of all nucleated cells at an optimal cut point of 0.64%. **E. Overall survival** stratified by stromal B cell percentage of all nucleated cells at an optimal cut point of 0.64%. **F. Quantitative features of B cell aggregates.** Y axis indicates distance between the center of the aggregate and tumor compartment; and size of circle indicates number of CD20+ B cells in the aggregate. “P” indicates presence of a pathologist-assessed lymphoid aggregate (PALA). **G. Logistic regression performance in predicting radiographic response**using the presence or absence of PALA (median AUC = 0.62), presence or absence of B cell aggregates by DBSCAN (median AUC = 0.79), or stromal B cell percentage (median AUC = 0.85) as a predictor. For each predictor, AUC values from 5-fold cross validation repeated 20 times are presented. A single association p-value for each factor was obtained using logistic regression. These analyses were conducted in the subset of patients with all three predictors available (n = 47).

### Automated detection of B cell aggregates by DBSCAN

In several cases with high stromal B cell counts, these cells were noted to appear in lymphoid aggregates of varying sizes (**Figure 3B).** As such, we tested whether the structural organization of B cells and other cell types were associated with ICI response.

First, a board-certified pathologist reviewed corresponding hematoxylin and eosin (H&E) -stained sections from 47 evaluable cases. Pathologist-assessed lymphoid aggregates (PALA) were identified in 14 of these 47 cases and were further classified by the presence or absence of an apparent germinal center. The percentage of stromal B cells measured by mIF was significantly higher in cases with PALA (median 4.95% vs. 0.01%, p<0.001). However, the presence of PALA was not associated with radiographic response, PFS, or OS.

To determine whether an automated method may be more effective in discriminating responders and non-responders, we leveraged a density-based clustering algorithm, DBSCAN,^20,21^ to automatically detect and quantify B cell aggregates using mIF data (STAR Methods). Based on visual inspection of a range of aggregates, an aggregate was defined as a density of 100 or more CD20^+^ B cells within a 76-micron radius occurring anywhere in the tissue section. Using DBSCAN, at least one B cell aggregate was identified in 20 out of 50 cases; 13 of the 20 were identified in specimens from sites other than lymph nodes. Among the 13 non-lymph node specimens, lymphoid aggregates contained a median of 186 B cells (IQR: 113 to 468; **Figure 3F**) and the median distance of a lymphoid aggregate to the tumor edge was 220 μm (IQR: 94 to 496 μm). In the 47 pathologist-evaluable cases, there was moderate agreement between DBSCAN identification of B cell aggregates and PALA (κ = 0.58). (**Supplementary Table 2**).

We then compared the performance of quantitative stromal B cell percentage, PALA and DBSCAN in predicting response to study therapy (**Figure 3G**) among the 47 pathologist-evaluable cases. The stromal B cell percentage among all nucleated cells We demonstrated the highest predictive performance with a median cross-validation AUC of 0.85, followed by DBSCAN with a median cross-validation AUC of 0.75. PALA had the poorest performance with a median cross-validation AUC of 0.62

### B cell neighborhood analysis

Finally, we analyzed the spatial distribution of other immune cell populations in relation to stromal B cells in responders and non-responders to elucidate potential functional relationships between immune cell types. For each sample containing B cells, we randomly selected 1000 immune cells (including all immune cells if total number of immune cells < 1000) from four major cell types (CD8^+^, CD4^+^, Treg, and macrophages) located within 250 μm of the nearest B cell and plotted their distances to the nearest B cell. In 34 non-lymph node samples containing B cells, CD4^+^ cells were found to be enriched in proximity to B cells, while CD68^+^ macrophages were relatively enriched at greater distances **(Figure 4A)**. The spatial proximity of CD4^+^ and CD8^+^ T cells to B cells was more pronounced in cases with radiographic response to ICI (n = 13), compared to those without (n = 21) **(Figure 4C**).

**FIGURE 4.**
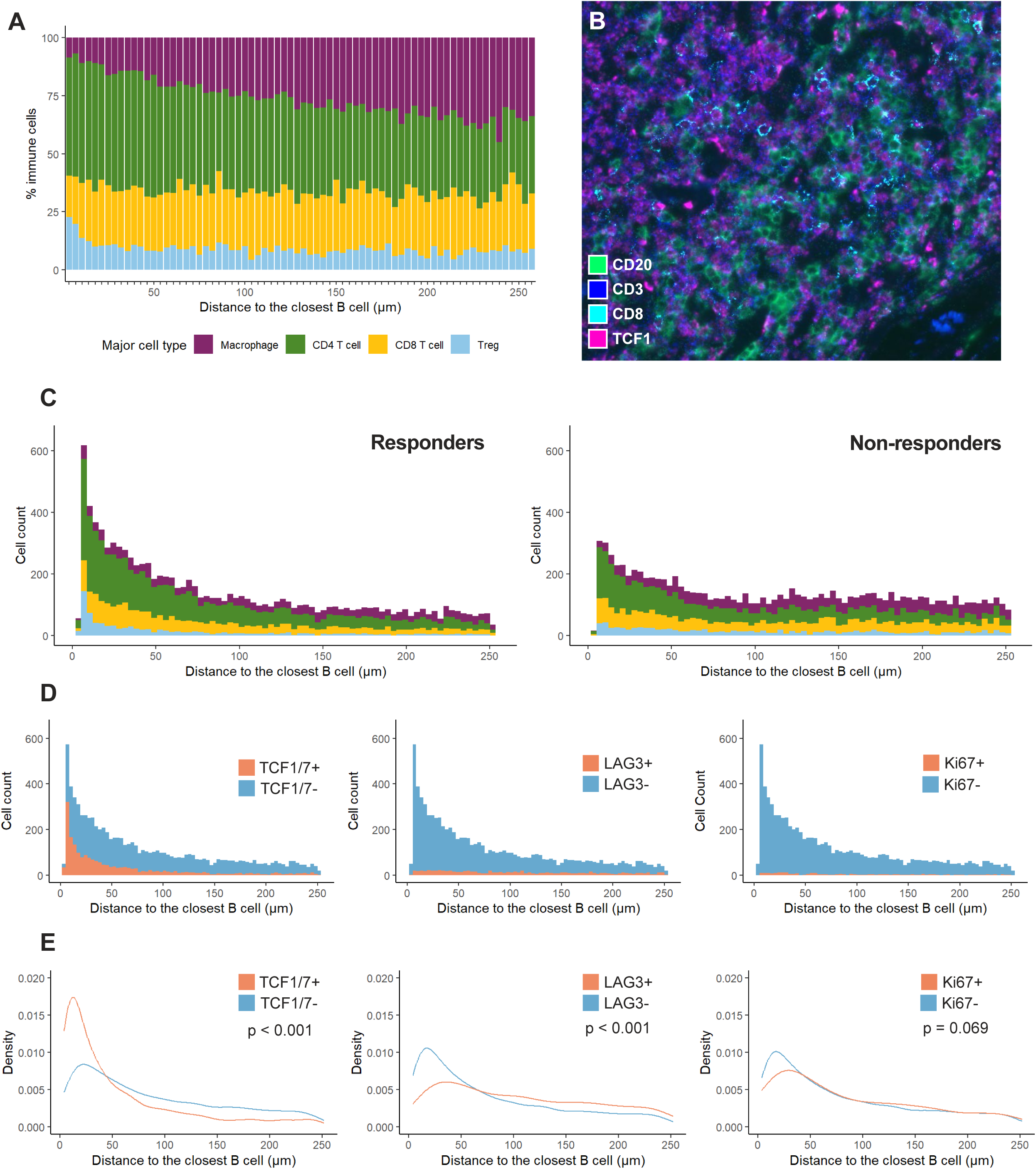
B cell spatial neighborhood analysis. **A. Spatial distribution of four immune cell types in B cell neighborhoods excluding lymph node samples (n = 34).** For each case, 1000 cells were randomly sampled within 250 μm from the nearest B cell and their relative proportions are plotted as a function of distance from the nearest B cell (μm). **B.** Representative image of TCF1**^+^**CD3**^+^** CD8**^-^** cells in proximity to a CD20+ B cells in the stroma of a soft tissue metastasis. **C. Spatial distribution of four immune cell types in relation to B cells in responders and non-responders.**Counts of 10,000 randomly sampled immune cells within 250 μm from the nearest B call among responders (n = 13) and non-responders (n = 21) as function of distance to the nearest B cell. **D.** Counts of TCF1**^+/-,^** LAG-3**^+/-^** and Ki67**^+/-^** CD3**^+^** T cell subsets as function of distance to the nearest B cell. **E. Densities of TCF1**^+/-^**, LAG-3**^+/-^ **and Ki67**^+/-^ **CD3**^+^ **T cell subsets as function of distance to the nearest B cell.** Densities for each cell type calculated as the number of cells at a given distance divided by all cells of that subtype within 250 μm of the nearest B cell. Spatial distributions compared using the **Kolmogorov-Smirnov test.**

We then examined functional subsets of CD3^+^ T cells in relation to B cells (**Figure 4D-E**). Among all CD3^+^ T cells, those expressing TCF1^+^ were more likely to be found near B cells compared to TCF1^-^ cells (p < 0.001). Additionally, LAG-3^-^ cells were more commonly located near B cells than LAG-3^+^ T cells (p < 0.001). There was no significant difference in the spatial distribution between Ki67^+^ and Ki67-T cells near B cells (p = 0.069).

## DISCUSSION

While the infiltration of CD8^+^ cytotoxic T cells has been considered one of the canonical indicators for response to ICI therapy, more recent studies have highlighted the role of other immune cell types in driving effective anti-tumor immunity in melanoma. For instance, tumor-infiltrating CD4^+^ cells have been found to react to HLA class II-restricted neoantigen^22^ and can eradicate major histocompatibility-deficient tumors that evade CD8+ T cell targeting.^23^

In this context, we performed a quantitative assessment of immune cells infiltrating pre-treatment tissue samples from patients with unresectable melanoma treated with ICIs. Using this approach, we identified a striking association between ICI response and the fraction of stromal B cells in pre-treatment melanoma samples. While the presence of PALA was not associated with ICI response, the quantitative assessment of the stromal B cell fraction and identification of B cell aggregates by DBSCAN effectively discriminated responders and non-responders. Interestingly, the quantitative percentage of B cells performed better than B cell aggregates discriminating responders and non-responders, suggesting that even more distributed stromal B cell infiltrates may play an important role in mediating anti-tumor immunity. These findings provide quantitative insights into the spatial features and the potential biologic relevance of B cell aggregates as potential drivers of response to ICI therapy in melanoma and highlight the potential of automated pathology workflows in the analysis of spatial data relevant to cancer immunotherapy.

This finding is consistent with prior reports regarding the potential significance of B cells in ICI response^24^— specifically, the presence of TLS has been associated with ICI response in melanoma,^15^ sarcoma,^17^ non-small cell lung cancer,^18^ and other tumor types.^16^ Although these data highlight the potential for TLS as a predictive biomarker of ICI therapy, it is important to note that TLS are complex three-dimensional structures initially described in a range of clinical settings including chronic infection^25^ and rejection of solid organ transplants.^26^ Unlike other tissue biomarkers studied in the context of immunotherapy, such as PD-L1 and TMB, TLS are comprised of multiple components, and there is no consensus definitions for these structures. For instance, the criteria for defining a TLS range from as few as 50 cells^16^ to as many as 700 cells, as seen in the PEMBROSARC trial.^17^ Also, while there is broad consensus that a surrounding T-cell infiltrate is important for defining a TLS,^27^ the specific markers used to identify these T cell components vary widely between studies. Perhaps most importantly, these definitions often require the specialized interpretation of histological information by a pathologist, raising questions about reproducibility of results.

Recognizing the potential for spatial biology tools to refine and standardize proposed TLS identification methods.^28^ We applied the spatial clustering algorithm, DBSCAN, to our data set to detect B cell aggregates using the CD20 channel output in mIF images. DBSCAN successfully identified B cell aggregates in a subset of cases and captured the majority of cases in which lymphoid aggregates were identified by a pathologist. However, DBSCAN-identified aggregates were more effective than PALA at discriminating responders and non-responders to ICI therapy. With further validation in larger data sets, automated pipelines like DBSCAN could significantly enhance TLS identification, either using CD20 immunohistochemistry, mIF-labeled, or clinical H&E stained slides.^29^ Application of this technique to larger tissue cohorts in melanoma and other tumors will be help optimize parameters that could be used to inform a digital clinical assay going forward. For instance, optimal thresholds for aggregate size, cell number, and/or distance from the tumor compartment could be identified to best distinguish ICI responders from non-responders in different tumor types.

Beyond standardizing TLS definitions, quantitative spatial analyses using mIF data can generate new hypotheses regarding the functional interplay between B cells and other immune cells within the tumor microenvironment. To address this, we leveraged additional markers from the mIF panel to identify functional T cell subsets proximal to B cell aggregates. Interestingly, TCF1^+^ T cells were significantly enriched in areas close to B cells; these TCF1^+^ cells may represent T follicular helper cells, which populate the T cell zones of mature TLS^30^ and rely on the expression of TCF1 for their differentiation^31,32^

TCF1 is also a marker for stem-like precursor exhausted T cells (T_pex_)^33^, which have been shown to expand following exposure to ICI therapy,^34,35^ and whose absence diminishes the response to ICI.^36^ Given the importance of these cells in mediating response to ICI therapy, it is possible that B cell aggregates form a spatial niche that promotes either the trafficking or persistence of these cells and bolster their ability to effect ICI-driven tumor killing. Other groups have similarly identified a spatial relationship between TLS and naïve T cells expressing TCF7 RNA transcripts in melanoma, which was also associated with a favorable response to immune checkpoint blockade.^37^ In other tumor types, an association with TCF1^+^ T cells and TLS has also been identified in oral squamous cell carcinoma and associated with favorable clinical outcomes in that patient population. Conversely, LAG-3-T cells were significantly enriched at farther distances from stromal B cells. While the potential relevance of this association to ICI response warrants further in-vitro, it is consistent with prior reports that canonical pro-inflammatory cytokines secreted by B cells, including TNF-alpha and IL-6, do not stimulate upregulation of LAG-3 on T cells.^38^

Taken together, spatially annotated cell-level mIF data has the potential to standardize identification of complex structures associated with ICI response and generate functional hypotheses about how these structures may interact with other cells in the tumor microenvironment. With further prospective validation, and in combination with other - omics (e.g., transcriptomics, epigenomics), quantitative spatial analysis of mIF/mIHC could guide the development of clinically relevant predictive biomarkers for selecting immunotherapies in unresectable melanoma and identify novel targets to further improve immunotherapy outcomes.

### Limitations of the study

Despite analyzing over 20 million individual cells, this study’s findings are based on a retrospective cohort from a single institution. Additionally, patients in this retrospective cohort received a variety of ICI regimens, and tissue was not collected at a uniform pre-treatment time point for all patients.

### Conclusion

Quantitative assessment of stromal B cells is strongly associated with response to ICI therapy in unresectable melanoma. Spatial analysis of cell-level mIF data allows for automated characterization of the size and distribution of B cell aggregates, and identification of spatial relationship between B cells and functional T cell subsets generate mechanistic hypotheses of how these features may influence anti-tumor immunity.

## Data Availability

All code describing cell phenotyping pipeline, application of DBSCAN for quantification of B cell aggregates, and B cell neighborhood analysis are available on GitHub.
Raw mIF data files and de-identified clinical data are available by request to the corresponding author.

## ACKNOWLEDGMENTS

This work was supported in part by the Memorial Sloan Kettering Cancer Center (MSK) NCI Core Grant P30 CA008748 and grant support from the V Foundation (T2021-007, M.K.C and R.S.).

## AUTHOR CONTRIBUTIONS

**J.W.S.** Writing—original draft, Supervision; **X.P.** Methodology, formal analysis; **F.D.E.** Methodology, Formal analysis, Data curation, Writing—review & editing; **A.P.M.** Investigation, Formal analysis; **M.Y.** Investigation; **C.M.** Project administration, Data curation; **N.A.** Investigation; **R.V.** Investigation; Writing—review & editing; **M.Z.** Investigation; **J.L.** Investigation, methodology; **M.B;** Writing--review & editing; **Y.L.** Investigation; **M.A.P.** Writing--review & editing, Resources; **T.J.H.** Conceptualization, Funding acquisition, Resources, Writing—review & editing; **K.S.P.** Supervision, Methodology, Writing—review & editing; **M.K.C.** Conceptualization, Funding acquisition, Writing—review & editing; **R.S.** Supervision, Methodology, Writing—review & editing.

## STAR METHODS

### RESOURCE AVAILABILITY

#### Lead contact

Further information and requests for resources should be directed to and will be fulfilled by the lead contact, James Smithy.

#### Materials availability

##### Data and code availability

All code describing cell phenotyping pipeline, application of DBSCAN for quantification of B cell aggregates, and B cell neighborhood analysis are available on GitHub.

Raw mIF data files and de-identified clinical data are available by request to the corresponding author.

### EXPERIMENTAL MODEL AND STUDY PARTICIPANT DETAILS

Patients with unresectable melanoma who were treated with immunotherapy at MSK and had pre-treatment tissue available were retrospectively identified. 27 patients were enrolled in clinical trials at MSK (protocols 09-155, 12-020, 12-179, 13-075, 13-169, 15-126, and 17-162), and an additional 23 patients were treated with immunotherapy as part of standard of care or expanded access (protocols 14-093 and 14-200). For these patients consent for analysis was obtained though protocols 12-245 and 06-107. All tissue analysis was performed through biospecimen protocol 19-114

### METHOD DETAILS

#### Assessment of clinical outcomes

Clinical outcomes were assessed for the first anti-PD-(L)1-based treatment line following tissue collection. For the four patients who did not receive anti-PD-(L)1-based treatment, the first line of treatment with any immune checkpoint inhibitor was examined. Radiographic response was assessed via RECIST v1.1,^39^ immune-related response criteria (irRC),^40^ or investigators’ clinical assessment. PFS and OS were calculated from the start of treatment.

#### Multiplex immunofluorescence (mIF) staining

Whole tissue sections were stained using Ultivue UltiMapper I/O Immuno8 Kit (Cambridge, MA, USA) containing CD8, PD-1, PD-L1, CD68, CD3, CD20, FoxP3, and pancytokeratin1+1SOX10 followed by opal tyramide staining containing TCF1, TOX, Ki67, LAG-3 as previously described.^41^ Whole slide imaging was acquired with Zeiss AXIO Scanner and tissue and cell segmentation were performed in using standard HALO workflows (Indica Labs, Albequerque, NM; Figure 1A-C)

#### Two-round staining with UltiMapper I/O Immuno8 Kit (Ultivue, ULT30801, Lot 204375)

Reagent assay and slide preparation were performed as the description of Ultivue ready to use protocol. FFPE tissue sections cut at 4-µm-thickness were baked for 1 h at 62 °C in vertical slide orientation prior to deparaffinization and 20 min of antigen retrieval (Bond ER2; Leica, AR9640) performed on a Leica Bond RX automated research stainer followed by eight DNA barcode:conjugated primary antibody cocktails (**Supplementary Table 3**) for a 20 min incubation. Once the sample had been incubated with the primary antibody cocktails, the DNA barcodes of each target were amplified to improve the sensitivity of the assay. Amplification was carried out simultaneously for all targets. Nuclear counterstain was applied after the amplification step (reagent in the UltiMapper I/O Immuno8 kit). Fluorescent probes complementary to the barcodes were then added to the sample, to bind and label each round of targets. Each target within a single imaging round was labeled with a spectrally distinct fluorophore to enable multiplexed whole slide imaging without the need for spectral unmixing. Using standard fluorescent filters: FITC, TRITC, CY5 and CY7, the four markers of the first-round staining were CD8, PD1, PD-L1 and CD68. The slides were mounted with ProLong Gold antifade reagent mounting medium (Invitrogen P36930). The full slide images were acquired using a Zeiss AXIO Scanner at 200X final magnification. After scan, the first-round stained slide was passively de-cover slipped by soaking the slide in PBS until the coverslip fell off, then the slide was washed with PBS (3×5 min) to remove the mounting medium, reloaded onto the BondRX strainer to start the second-round staining with Exchange Initiator (Ultivue, ULT00133) incubation 3X10 min, followed by the fluorescent probe cocktails which contained Exchange Neutralizer (ULT00131) and second set of four DNA barcode conjugated antibodies (CD3, CD20, FOXP3, panCK+SOX10) for 12 min incubation. The slides were mounted as described for first-round staining and new whole slide images acquired. We utilized a panCK+SOX10 cocktail as part of the UltiMapper I/O Immuno8 kit to identify cells with either an epithelial or neural crest/melanocytic derivation. To compensate for combination staining within this channel, the resulting stained images were individually analyzed with areas of melanoma tumor morphology identified assisted by panCK+SOX10.

#### Four-color multiplex opal tyramide staining

Coverslips were removed after the round 2 scan as described above. The sections were thoroughly washed with PBS and slides reloaded into the BondRX Stainer. Four-color multiplex staining was performed with validated primary antibodies in optimal concentration and stripping condition which were selected by MSK pathologist (TH) according to the results of diaminobenzidine IHC staining on the Leica BondRX Stainer. Four sequential cycles of staining, each round of which included a 10 min blocking (Akoya antibody diluent/block ARD1001) and 30 min primary antibody incubation. The primary antibody detection was performed using HRP conjugated species-specific secondary with 10 min incubation. The HRP-conjugated secondary antibody polymer was detected using fluorescent tyramide signal amplification using Opal dyes 520, 570, 650 and 780 (**Supplementary Table 4**) After each staining cycle, two heat-induced stripping steps of the primary/secondary antibody complex were performed using Akoya AR9 buffer (AR900250ML) and Leica Bond ER2 (90% AR9 and 10% ER2) at 100 °C for 20 min, 3x 2 min Bond washing steps preceded the next cycle. After 4 sequential rounds of staining, nuclear counterstaining was performed with Hoechst (Invitrogen 33342) and mounted with ProLong Gold antifade reagent mounting medium. Whole slide imaging was acquired with Zeiss AXIO Scanner.

The images from three rounds were stacked with Ultivue Ultistacker v1.0b6 software to get the final 12-marker stacked images which can be visible and analyzable in HALO software (Indica Labs, Albequerque, NM). All markers were validated by assessing sensitivity correlations to monoplex DAB, including following the stripping process.

#### Multiplex cell annotation

Stacked TIFF images were reviewed and analyzed in HALO. Areas of necrotic or folded tissue were manually excluded, and representative stroma, tumor, and glass regions were manually selected to inform a regional classifier. The AI module was used to perform nuclear segmentation based on the Hoechst stained mask in the OPAL protocol. Whole cells were defined by dilating the nuclear segmentation by 1.2 microns beyond the Hoechst signal, and each cell was assigned a unique ID.

#### Slide segmentation & cell phenotype assignment

For every image, individual markers were thresholded using the mean intensity via the HALO HighPlex FL 4.1.3 module. This process separates positive cells from non-specific staining based on a visually determined cutoff. Subsequently, all HALO outputs were imported into R (v4.1.1) for further analysis, which included assigning positive/negative labels to cells for each marker per image, as well as compiling the mean intensity values for all cells.

Inspired by the methodology described by Zhang et al for each individual cell,^19^ the raw intensity value *X_m_* of marker m was scaled to probabilities *EP*(*X_m_*) ∈ [0,1], defined as

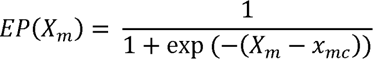

Where *x_mc_* is the human-set threshold for marker m in the image. Both *X_m_* and *x_mC_* were transformed using asinh() to mitigate the effect of a long tight tail. Here, *EP*(*X_m_*) = *P*(*m*^+^) =1 - *P*(*m*^−^) represents the probability of the marker m being positive for the cell. A value of *EP*(*X_m_*) significantly great than 0.5 indicates confidence in the marker’s positivity; conversely, a value significantly less than 0.5 indicates its negativity. When *EP*(*X_m_*) is approximately 0.5, additional information from other channels may be requires for better categorizing cell types.

Our cell phenotyping pipeline comprises two main steps: 1) phenotyping major cell types and 2) identifying functional cell subsets. Initially, seven major cell types—tumor/epithelium, macrophage, T_reg_, CD8^+^ T cell, CD4^+^ T cell, B cell, and others—were defined based on their expression of six lineage markers: SOX10/CK, CD8, CD3, CD20, CD68, FoxP3 (**Supplementary Table 5**). The sum of the macrophage, T_reg_, CD8^+^ T cell, CD4^+^ T cell, B cell populations was used to reflect all immune cells for each case. For each cell, we computed a similarity score for each major cell type. For instance, the similarity score for a cell being a CD8 T cell is calculated as

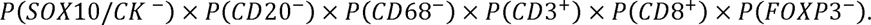

During cell phenotyping, cells were assigned to the major cell type with the highest similarity score. This approach utilizes both the human-set threshold and the absolute cell-level signal intensity from all lineage markers, enhancing the robustness of our phenotyping results against noise in multiplexed immunofluorescence images. For identifying functional cell subsets, markers TCF1, Ki67, TOX, LAG-3, PD-1, and PD-L1 were integrated with lineage assignments to define functional cell populations.

#### Identification of B cell aggregates by DBSCAN

After classifying and counting the B cells in the Multiplex IF (mIF) slides, the coordinates of the B cells were used to detect the B cell aggregates. The B cell aggregates were defined as dense regions where the density of the B cells is high and clustered together. By visually inspecting the distribution of B cells in the mIF slides, if the number of B cells is higher than 100 cells within a circle with a radius of 75 µm, it is defined as an aggregate of B cells.

To computationally detect the B cell aggregates in the mIF slides, we incorporated and modified the DBSCAN (Density-Based Spatial Clustering of Applications with Noise) algorithm.^21^ DBSCAN is a clustering method used in data mining and machine learning applications in large datasets. Since the number of cells in the whole mIF slides within this study is high, and the number of aggregates within each slide is unknown, the DBSCAN algorithm has a significant advantage over other traditional clustering methods such as K-means where a prior number of clusters is required. The coordinates of the B cells from the detected aggregates were then used to analyze the morphological features as well as the spatial distribution of the B cell aggregates within the tumor. To investigate the morphological features of the detected B cell aggregates, a convex hull was fit to the B cells (connecting the outermost cells) and the area, roundness, radius, skewness, and density of cells (number of cells per unit of area) were calculated and studied.

The spatial distribution of the B cell aggregates was studied, and the distance of each B cell aggregate from the tumor was calculated. To computationally calculate the distance of the B cell aggregates from the tumor, we detected the centroid of the aggregates. Then, the distance between the centroid of the aggregate and the tumor region was calculated for each cluster. Finally, the radius of the aggregate was deducted from the calculated distance between the centroid of the aggregate and the tumor region. The smaller the distance, the closer the aggregate is to the tumor. If the distance value is negative, it indicates that the B cell aggregate is located inside the tumor region.

### QUANTIFICATION AND STATISTICAL ANALYSIS

To examine cell type composition, 265 cell population percentages and five hypothesis-driven cell population ratios were calculated within each of the three tissue compartments (stroma, tumor, and stroma within 100 μm of the nearest tumor cell). Specifically, counts of each major cell type, all possible subsets expressing up to two functional markers, and total immune cells were expressed as percentages of all cells in the compartment. In addition, counts of each immune major cell type and all possible subsets expressing up to two functional markers were expressed as percentages of all immune cells in the compartment. The following ratios were calculated: CD4 T cells to CD8 T cells, macrophages to CD3 T cells, TCF1+ CD4 T cells to TOX+ CD4 T cells, TCF1+ CD8 T cells to TOX+ CD8 T cells, and TCF1+ immune cells to TOX+ immune cells. Population percentages were illustrated using heatmaps and bar plots. Within each compartment, the Wilcoxon rank sum test with false discovery rate correction was used across all percentages and ratios to identify cell populations enriched or depleted in responders relative to non-responders.

Area under the receiver operating characteristic curve (AUC) was used to quantify the ability of stromal B cell percentage to discriminate responders and non-responders. Youden’s J statistic was then used to identify an optimal cut point in stromal B cell percentage and classify patients into two groups. Associations between stromal B cell classification and survival outcomes (PFS and OS) were illustrated using Kaplan-Meier curves and assessed using the log-rank test. These analyses were repeated excluding patients with lymph node metastases. Associations between stromal B cell classification and patient characteristics were assessed using Fisher’s exact test and Pearson’s Chi-squared test.

The association between pathologist-assessed lymphoid aggregates (PALA) and stromal B cell percentage was assessed using the Wilcoxon rank sum test. Characteristics of B cell aggregates identified by DBSCAN in non-lymph node samples were summarized in a dot plot. Agreement between pathologist identification of lymphoid aggregates and DBSCAN identification of B cell aggregates was assessed using Cohen’s kappa. To compare performance in predicting response across stromal B cell percentage, DBSCAN, and PALA, logistic regression with response as the outcome was performed among the subset of patients with all three factors available. Stromal B cell percentage was folded root transformed for logistic regression. Using each factor as a predictor, test AUC values were calculated via 5-fold cross validation repeated 20 times, illustrated using box plots, and summarized by median. A single association p-value for each factor was obtained via logistic regression on the entire subset of patients with all three factors available.

Distances between immune cells and the nearest B cell were illustrated using bar plots, histograms, and density plots. Distributions of distance to the nearest B cell were compared between cell types using the Kolmogorov-Smirnov test.

Two-sided p values lower than 0.05 were considered statistically significant. All analyses were conducted using R version 4.3.2.

## SUPPLEMENTAL INFORMATION

**Supplementary Table 1.**
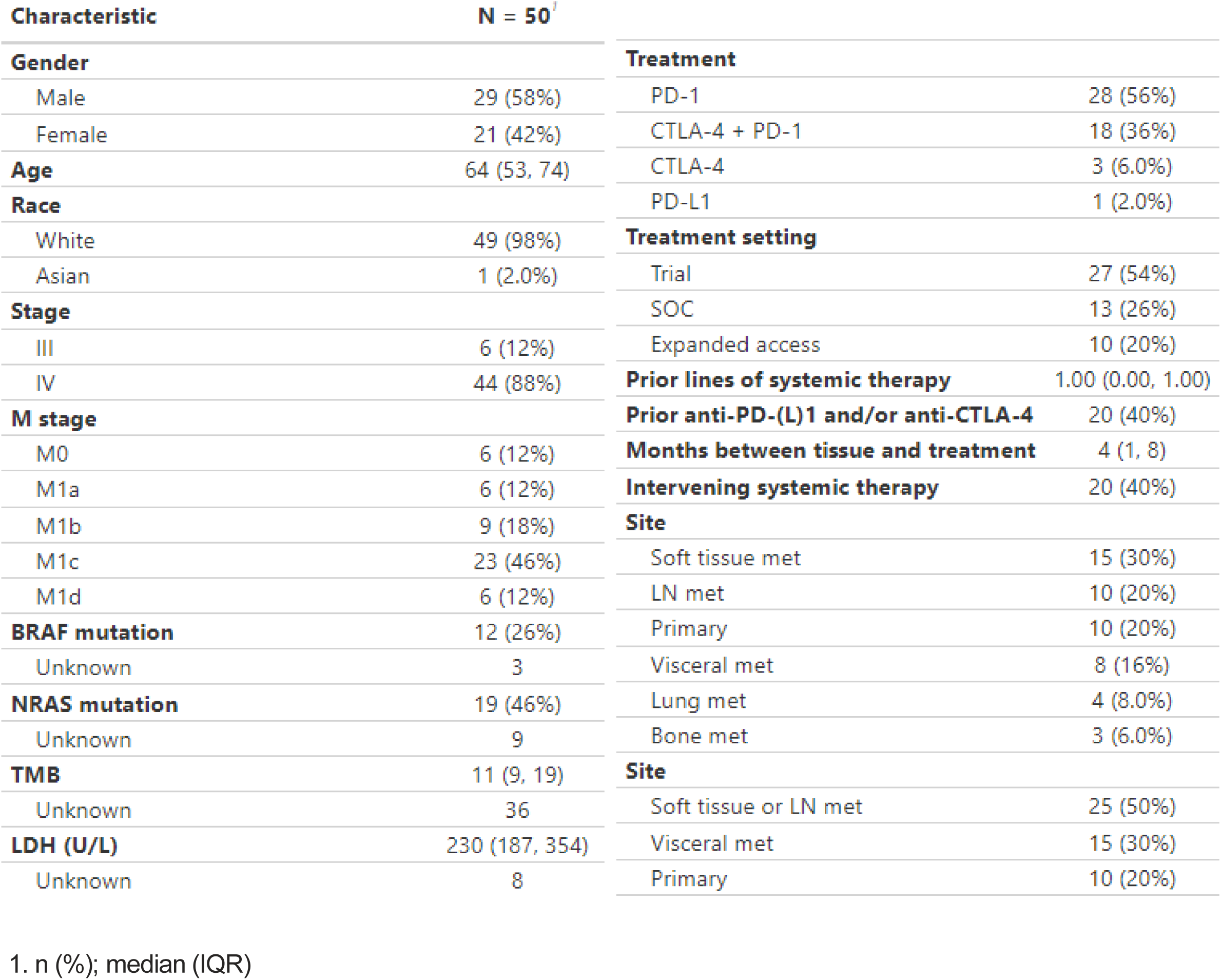
Patient characteristics.

**Supplementary Table 2.** Concordance of PALA and the presence of B cell aggregates by DBSCAN (n = 47)

|  | DBSCAN+ | DBSCAN- |
| --- | --- | --- |
| PALA+ | 12 | 2 |
| PALA- | 7 | 26 |

$\kappa = 0.58$

**Supplementary Table 3.**
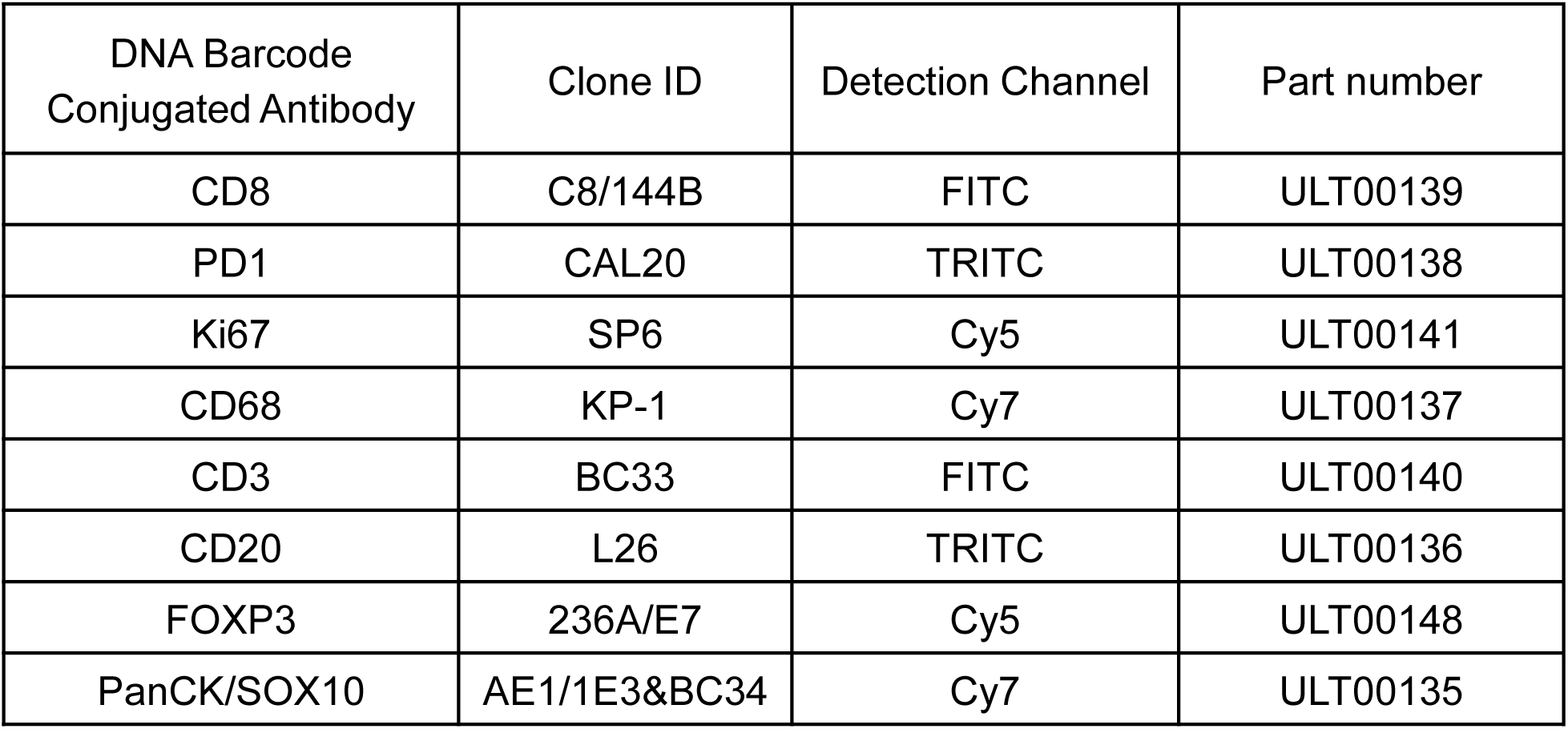
DNA-barcode conjugated antibody details.

**Supplementary Table 4.** Four-color multiplex opal tyramide antibody details.

| Primary Antibodies | Antibody Clone | Vendor | Titration | Secondary Antibodies | Detection Dye (cycle) |
| --- | --- | --- | --- | --- | --- |
| TOX | E6I3QBF | CST | 0.644µg/ml | Invitrogen Rabbit Poly HRP (B40962) | Opal 520 (1)<br>1:100 |
| LAG3 | D2G40 | CST | 7.8µg/mL | Invitrogen Rabbit Poly HRP (B40962) | Opal 570 (2)<br>1:150 |
| PD-L1 | 73-10 | Abcam | 0.18ug/ml | Invitrogen Rabbit Poly HRP (B40962) | Opal 650 (3)<br>1:300 |
| TCF1/7 | C63D9 | CST | 1.33µg/ml | Invitrogen Rabbit Poly HRP (B40962) | Opal 780 (4)<br>1:100 |

**Supplementary Table 5.** Assignment of major cell types based on combinatorial expression of lineage markers.

| Cell type | panCK<br>/SOX10 | CD20 | CD68 | CD3 | CD8 | FOXP3 |
| --- | --- | --- | --- | --- | --- | --- |
| Tumor v.<br>Epithelium | 1 | 0 | 0 | 0 | 0 | 0 |
| Macrophage | 0 | 0 | 1 | 0 | 0 | 0 |
| B cell | 0 | 1 | 0 | 0 | 0 | 0 |
| CD8<sup>+</sup> T | 0 | 0 | 0 | 1 | 1 | 0 |
| CD4<sup>+</sup> T | 0 | 0 | 0 | 1 | 0 | 0 |
| T<sub>reg</sub> | 0 | 0 | 0 | 1 | 0 | 1 |
| Other | 0 | 0 | 0 | 0 | 0 | 0 |

**Supplementary Figure 1.**
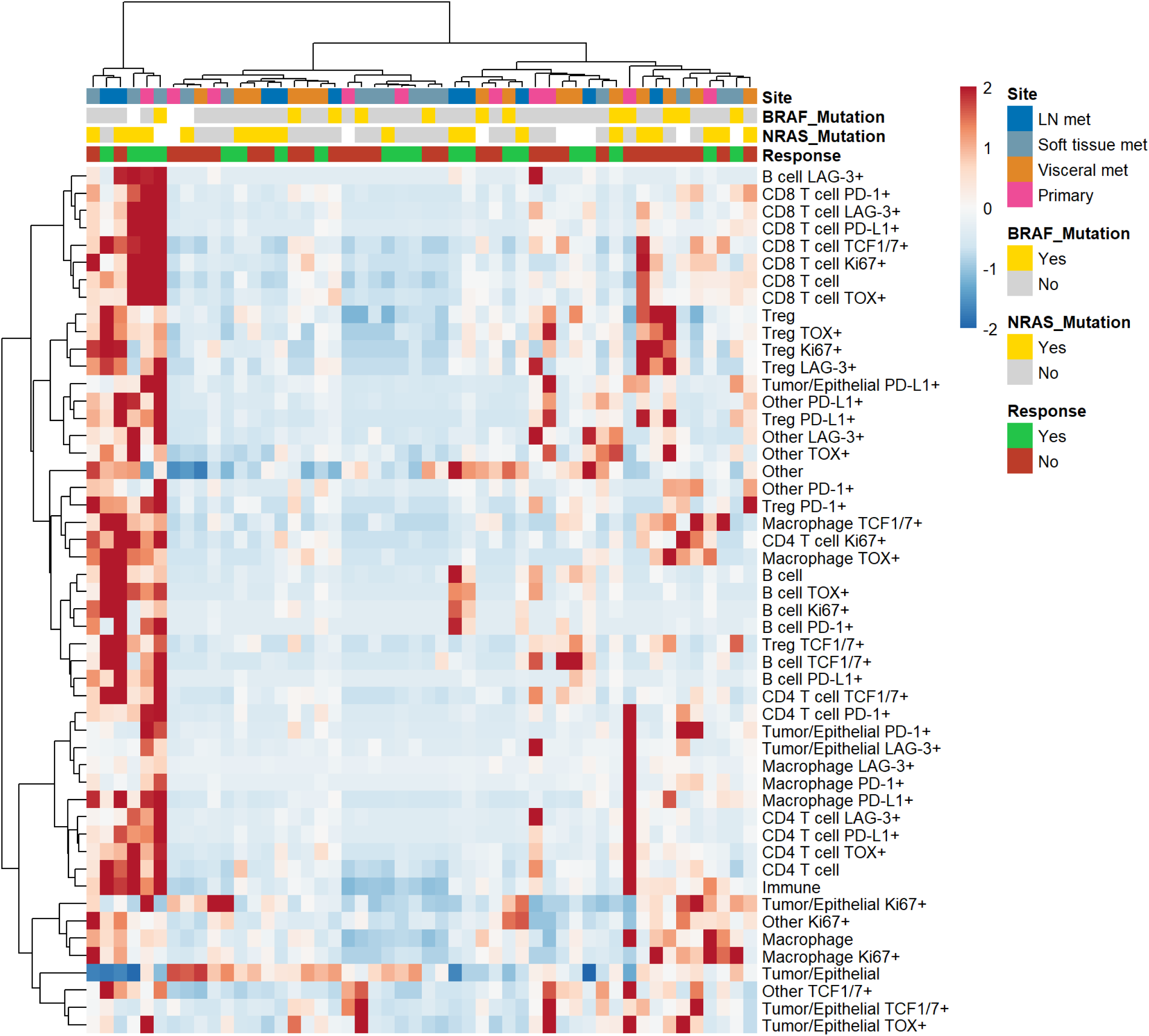
Heatmap of intratumoral percentages of cell populations and clinical characteristics. A subset of the 265 cell populations is presented for visibility: major cell types, each major cell type expressing each functional marker, and total immune cells.

**Supplementary Figure 2.**
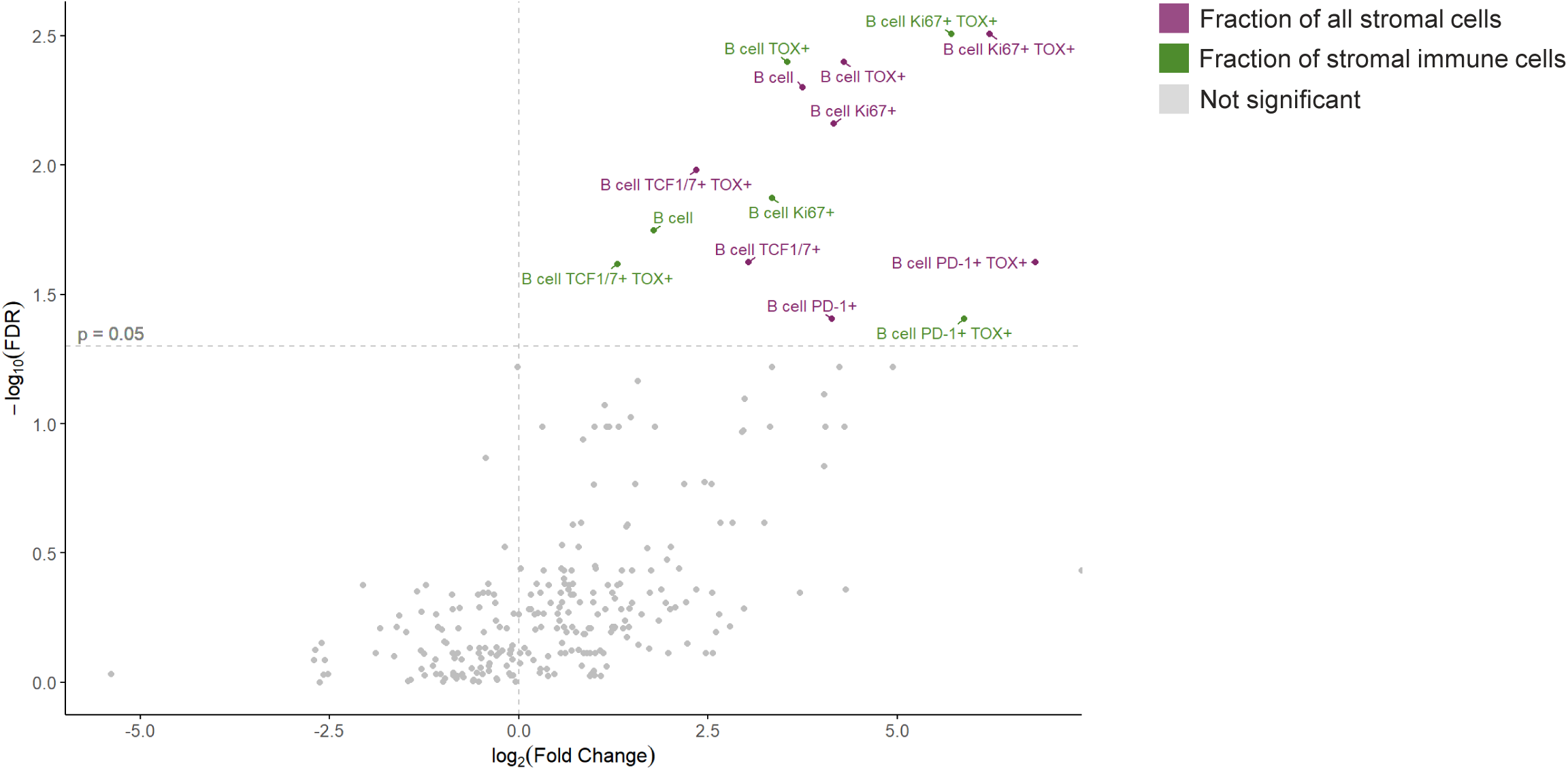
Volcano plot of stromal immune cell populations enriched in patients with complete or partial response to ICI therapy, limited to stroma within 100 μm of the nearest tumor cell. Immune cell populations were calculated as a percentage of all stromal cells (purple) and as a fraction of stromal immune cells (green).

**Supplementary Figure 3.**
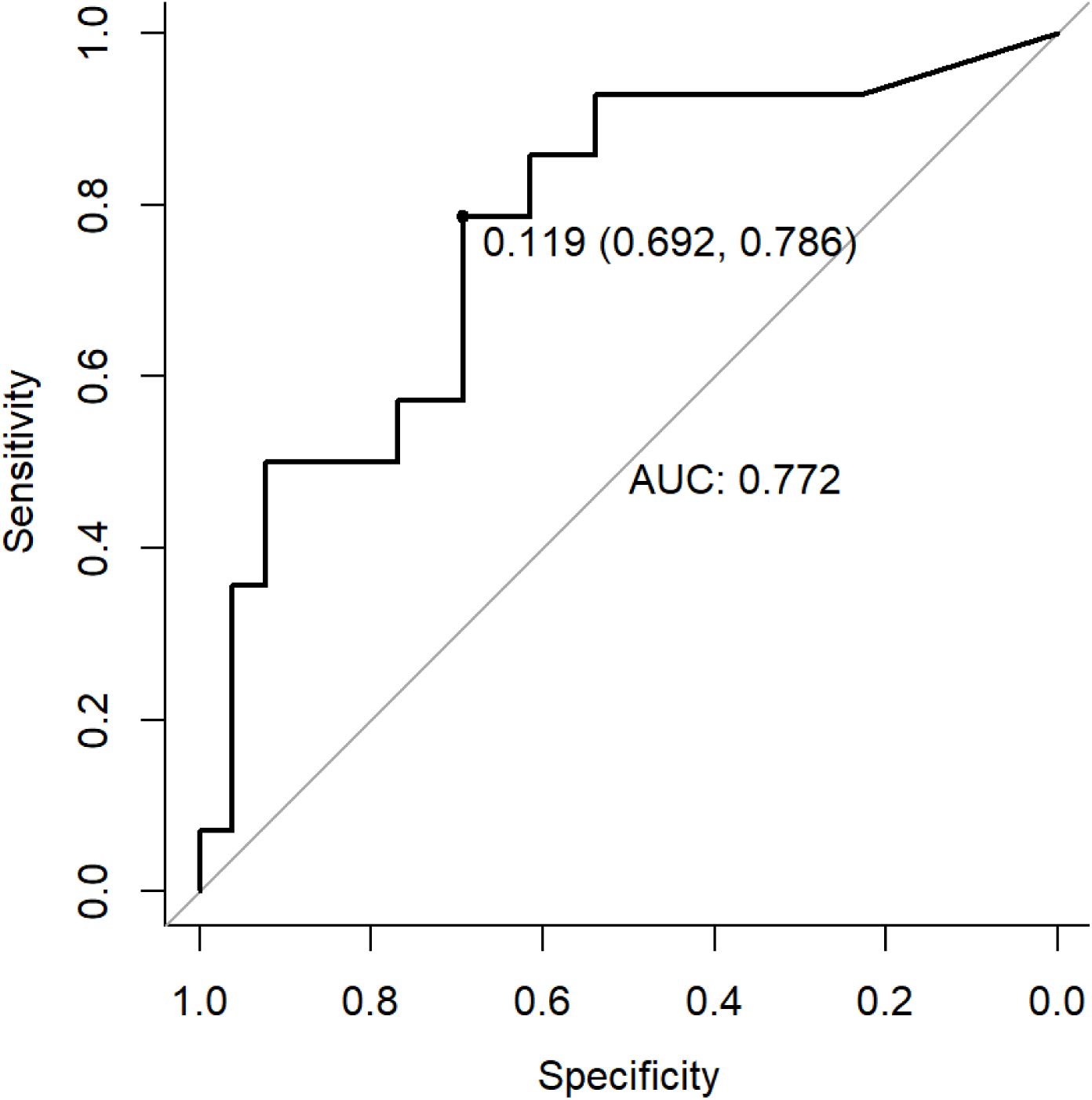
Receiver operating characteristic curve of. stromal B cell percentage as a predictor of radiographic response to ICI therapy, excluding lymph node metastases (n = 40). Area under the curve = 0.772; optimal cutpoint = 0.119%

